# THE VARIATION OF GENOME SITES ASSOCIATED WITH SEVERE COVID-19 ACROSS POPULATIONS: THE WORLDWIDE AND NATIONAL PATTERNS

**DOI:** 10.1101/2020.11.22.20236414

**Authors:** Oleg Balanovsky, Valerie Petrushenko, Karin Mirzaev, Sherzod Abdullaev, Igor Gorin, Denis Chernevskiy, Anastasiya Agdzhoyan, Elena Balanovska, Kryukov Alexander, Dmitriy Sychev

## Abstract

**Background:** The knowledge of clinically relevant markers distribution might become a useful tool in COVID-19 therapy using personalized approach in the lack of unified recommendations for COVID-19 patients management during pandemic. We aimed to identify the frequencies and distribution patterns of rs11385942 and rs657152 polymorphic markers, associated with severe COVID-19, among populations of the world, as well at the national level within Russia. The study was also dedicated to reveal whether population frequencies of both polymorphic markers are associated with COVID-19 cases, recovery and death rates.

**Methods:** We genotyped 1883 samples from 91 ethnic populations from Russia and neighboring countries by rs11385942 and rs657152 markers. Local populations which were geographically close and genetically similar were pooled into 28 larger groups. In the similar way we compiled a dataset on the other regions of the globe using genotypes extracted or imputed from the available datasets (32 populations worldwide). The differences in alleles frequencies between groups were estimated and the frequency distribution geographic maps have been constructed. We run the correlation analysis of both markers frequencies in various populations with the COVID-19 epidemiological data on the same populations.

**Findings:** The cartographic analysis revealed that distribution of rs11385942 follows the West Eurasian pattern: it is frequent in Europeans, West Asians, and particularly in South Asians but rare or absent in all other parts of the globe. Notably, there is no abrupt changes in frequency across Eurasia but the clinal variation instead. The distribution of rs657152 is more homogeneous. Higher population frequencies of both risk alleles correlated positively with the death rate. For the rs11385942 we can state the tendency only (r=0,13, p=0.65), while for rs657152 the correlation was significantly high (r=0,59, p=0,02). These reasonable correlations were obtained on the Russian dataset, but not on the world dataset.

**Interpretation:** Using epidemiological statistics on Russia and neighboring countries we revealed the evident correlation of the risk alleles frequencies with the death rate from COVID-19. The lack of such correlations at the world level should be attributed to the differences in the ways epidemiological data have been counted in different countries. So that, we believe that genetic differences between populations make small but real contribution into the heterogeneity of the pandemic worldwide. New studies on the correlations between COVID-19 recovery/mortality rates and population’s gene pool are urgently needed.

## Introduction

A novel coronavirus known as Severe Acute Respiratory Syndrome Coronavirus 2 (SARS-CoV-2) was first identified in China in December 2019 and due to its fast spread in three months it was identified as a pandemic by the World Health Organization [1]. The pathogen causes disease called coronavirus disease 2019 (COVID-19).

The disease course varies drastically among patients, from asymptomatic infection to death from the respiratory failure. It was reported that patients with severe COVID-19 were older and/or with a greater number of comorbid conditions [2]. Epidemiological indicators show that morbidity and mortality is highly variable not only between individuals but also among countries. So, to the date of paper writing the rate of COVID-19 cases in North European countries (Denmark, Estonia, Finland, Norway) was relatively low, while southern countries have experienced higher morbidity and mortality rates (Italy, Spain, and France) [3]. This can result from different social-demographic factors, environment factors (see [4]–[6] for correlation with the vitamin D levels) and, importantly, genetic factors. The genome-wide association study replicated on Italian and Spanish patients identified two key genome regions caused severe respiratory failure in COVID-19 patients [7]. The first site, *rs11385942* (insertion-deletion G>GA) is located within a cluster of six genes, whose functions are relevant to COVID-19. Notably, the association is evident for the entire haplotype block of 50kb length. So high LD in this site, unusual for this segment of chromosome 3, resulted from an evolutionary recent introduction of this haplotype into human gene pool from Neanderthals [8]. The second site, demonstrating less pronounced association with severe Covid-19 (*rs657152*) is located in the gene encoding ABO blood groups, which may also determine the severity of respiratory failure in COVID-19 patients. The paper presenting both associated sites [7] stated that their frequencies vary among 1000 Genomes project populations, but has not described the pattern of their variation.Thus, in our study we focus on the global population variation of both genomic sites, associated with the severe Covid-19.

We analyzed the variation of both SNPs in worldwide populations, based on this study dataset (1883 individuals) from North Eurasian countries and on the database compiled from the published data on other regions of the globe (3088 individuals). We aimed to unravel the patterns of geographic variation of these polymorphisms and investigate whether these genetic factors contributed to the variety of COVID-19 epidemiological parameters between different populations. In the lack of unified recommendations for COVID-19 patients’ management the knowledge of clinically relevant markers distribution may become a useful tool in COVID-19 therapy using personalized approach.

Therefore, the initial goal of this study was to identify the frequency of *rs11385942* and *rs657152* markers among populations at the global level as well as at the national level within Russia. The latter level is important due to the high ethnic heterogeneity of populations from Russia and neighbor states. The second goal was to determine whether there is any connection between the population frequencies of *rs11385942* and *rs657152* polymorphic markers and COVID-19 cases, recovery and deaths rates.

## Materials and methods

### Generating the dataset on the Russian and neighboring populations

The samples from populations of Russia and neighbor North Eurasian counties (former USSR and Mongolia) were received from Biobank of North Eurasia [9]. All sample donors gave the written informed consent, approved by the Ethical Committee of the Research Centre for Medical Genetics. The set of analyzed samples covers the large area of Russian and neighboring countries and includes most of ethnic groups from this area (Figure 1). The sampling strategy (similar to the one of the 1000 Genomes project) was focused on the indigenous populations; for example in East Siberia we sampled Yakuts and Evenks rather than ethnic Russians who migrated to Siberia within last 1-3 centuries.

**Figure 1.**
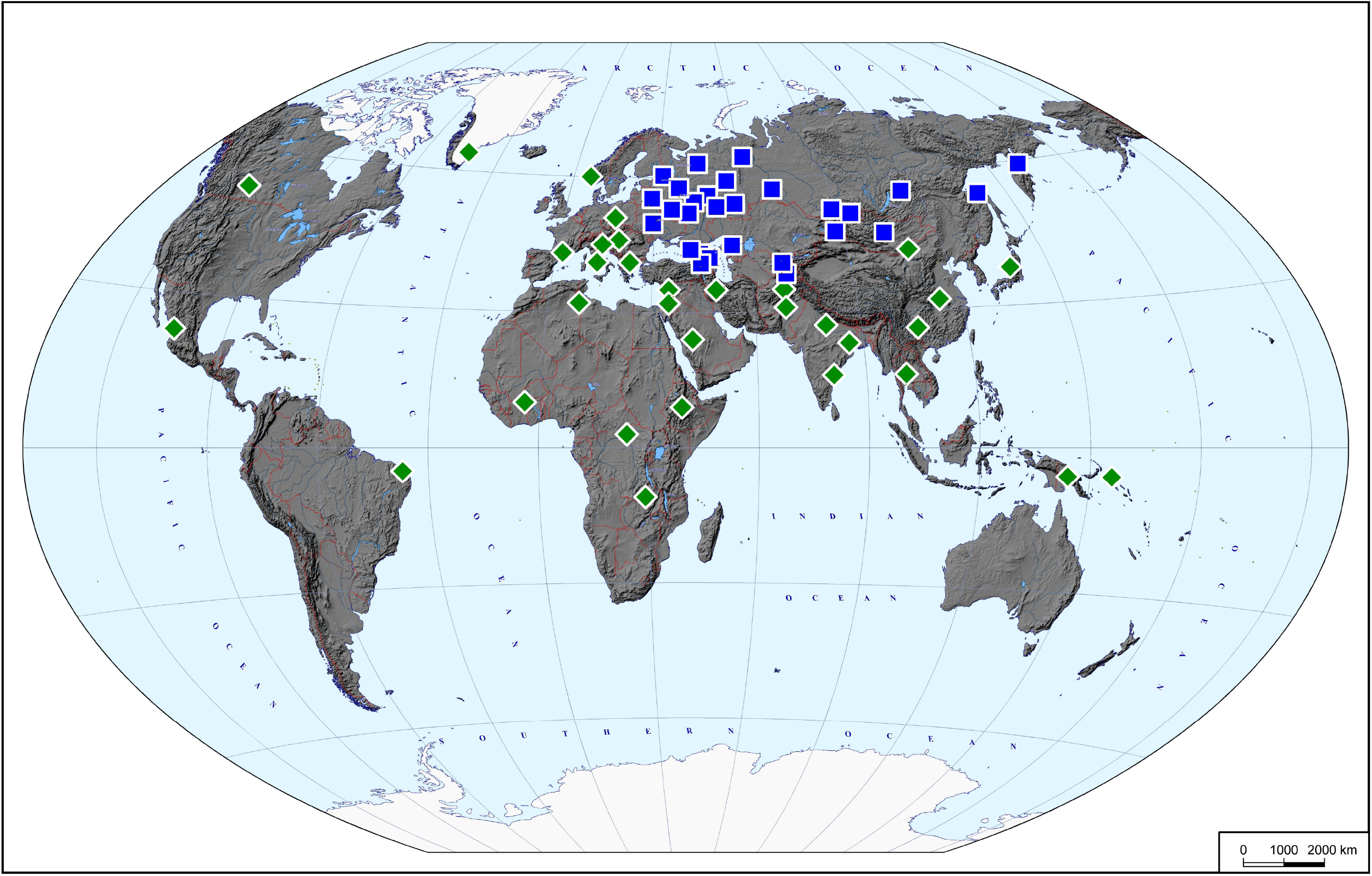
The studied populations. Blue squares designate the locations of the samples genotyped in this study (the dataset on Russia and neighboring countries). Green diamonds designate the locations of the samples genotyped in other papers and reanalyzed in this study (the world dataset).

In total, we genotyped 1883 samples from 91 ethnic populations from Armenia, Azerbaijan, Belorussia, Georgia, Kazakhstan, Kirgizia, Lithuania, Mongolia, Russia, Tajikistan, Ukraine, and Uzbekistan. We focused on the two markers, *rs11385942* and *rs657152*, associated with the severe Covid-19 [7]. We genotyped these samples by Infinium Omni5Exome-4 v1.3 BeadChip Kit (Illumina, USA) which includes 4,5 million SNPs. For this study we extracted genotypes of *rs657152* and genotypes of 250 SNPs located in the vicinity of *rs11385942* (positions 45700000-45900000 on the chromosome 3, Human Genome Build 37).

Most populations were represented by few samples each, while we needed sample size about 100 chromosomes per population to estimate the frequencies of the COVID-associated markers. Thus, local populations which were geographically close and genetically similar were pooled into larger groups. This pooling was made based on the genetic relationship between populations identified by means of the entire dataset of 4,5 million SNPs. The pooling procedure included i) standard filtering in PLINK [10] (geno 0.05, maf 0.01, mind 0.1, pruning indep-pairwise 1500 150 0.2), ii) running the set of PCAs by smartpca software [11], [12], and iii) identifying groups of populations with homogeneity within the groups but pronounced variation between the groups; we omitted some populations which could not stand alone because of the low sample size and could not be pooled because of their genetic peculiarities; the details of this pooling procedure are described in [13]. The pooling resulted in the final dataset of 1785 individuals from 28 pooled populations with the average sample size 128 chromosomes (64 individuals). Table 1 presents the list of these populations (“Russian dataset” section). Their geographic locations are present on the map (Figure 1).

**Table 1.** Frequencies of the two genetic markers associated with severe Covid-19 in various populations (“Russian” and “world” datasets). Note: samples sizes are given in chromosomes (twofold number of individuals).

| Region | Population | Country | Sample size | Latitude | Longitude | Frequency of *rs11385942_GA* | Frequency of *rs657152_A* |
| --- | --- | --- | --- | --- | --- | --- | --- |
| **Russian dataset (populations from Russia and neighboring countries)** |  |  |  |  |  |  |  |
| East European plain | Russians northern | Russia | 166 | 57,70 | 37,64 | 0,06 | 0,40 |
|  | Russians south-eastern | Russia | 102 | 52,12 | 41,02 | 0,10 | 0,41 |
|  | Russians south-western | Russia | 100 | 52,92 | 35,41 | 0,15 | 0,39 |
|  | Russians northernmost | Russia | 140 | 63,12 | 44,43 | 0,06 | 0,42 |
|  | Ukrainians | Ukraine | 158 | 49,83 | 29,42 | 0,16 | 0,51 |
|  | Komi and Udmurts | Russia | 168 | 59,23 | 54,35 | 0,15 | 0,40 |
|  | Chuvash and Mari | Russia | 106 | 56,05 | 47,47 | 0,10 | 0,46 |
|  | Mordovians | Russia | 80 | 54,59 | 42,77 | 0,05 | 0,38 |
|  | Tatars | Russia | 104 | 53,49 | 50,28 | 0,08 | 0,57 |
|  | Bashkirs | Russia | 86 | 54,15 | 56,45 | 0,12 | 0,42 |
|  | Belorussians and north-west Russians | Belorussia, Russia | 60 | 55,32 | 28,57 | 0,13 | 0,33 |
|  | Karelians and Veps | Russia | 118 | 60,42 | 31,92 | 0,14 | 0,36 |
| Siberia | Altaians | Russia | 154 | 52,43 | 88,43 | 0,04 | 0,48 |
|  | Tuvinians and Tofalars | Russia | 110 | 51,44 | 94,38 | 0,04 | 0,38 |
|  | Buryats, Khamnegans, and Yakuts | Russia | 114 | 55,74 | 114,28 | 0,04 | 0,42 |
|  | Chukchi, Koryaks, and Itelmen | Russia | 134 | 59,23 | 162,08 | 0,01 | 0,42 |
|  | Far East (Nanays, Ulcji, Nivkh, Evens) | Russia | 168 | 54,03 | 140,02 | 0,02 | 0,38 |
|  | Khanty, Mansi, and Nenets | Russia | 106 | 64,52 | 61,40 | 0,02 | 0,38 |
|  | SiberianTatars | Russia | 136 | 57,22 | 70,03 | 0,02 | 0,41 |
| Caucasus | West Caucasus | Russia | 174 | 43,89 | 41,47 | 0,11 | 0,27 |
|  | East Caucasus (Dagestan) | Russia | 158 | 42,07 | 47,36 | 0,11 | 0,52 |
|  | Central Caucasus | Russia | 128 | 43,04 | 44,50 | 0,05 | 0,42 |
|  | Transcaucasia (South Caucasus) | Armenia, Azerbaijan, | 154 | 40,83 | 44,65 | 0,14 | 0,47 |
|  |  | Georgia |  |  |  |  |  |
| Central Asia | Mongols Khalkh | Mongolia | 98 | 46,93 | 103,23 | 0,01 | 0,32 |
|  | Mongols (non-Khalkh) and Kalmyks | Mongolia, Russia | 156 | 47,56 | 87,74 | 0,04 | 0,47 |
|  | Kazakhs, Karakalpaks, Uigurs, and Nogais | Kazakhstan,<br>Uzbekistan, Russia | 88 | 44,91 | 54,46 | 0,01 | 0,45 |
|  | Uzbeks, Turkmens, and Kirghiz | Uzbekistan,<br>Kirgizia | 160 | 40,83 | 69,42 | 0,04 | 0,51 |
|  | Tajiks, Pomiri, and Yaghnobi | Tajikistan | 144 | 38,43 | 70,22 | 0,15 | 0,46 |
| World dataset (other countries) |  |  |  |  |  |  |  |
| South Asia | East_Indians | India | 90 | 23,00 | 86,00 | 0,23 | 0,47 |
|  | North_Indians | India | 186 | 27,00 | 80,00 | 0,29 | 0,51 |
|  | South_Indians | India | 96 | 16,00 | 81,00 | 0,22 | 0,40 |
|  | Afghanisthan | Afghanisthan | 64 | 35,00 | 69,00 | 0,11 | 0,44 |
|  | Pakistanis | Pakistan | 336 | 31,00 | 69,00 | 0,15 | 0,45 |
| Europe | Franch, Spaniards, Basque | France, Spain | 184 | 43,00 | 2,00 | 0,07 | 0,32 |
|  | Italians | Italy | 88 | 41,00 | 13,00 | 0,13 | 0,35 |
|  | North Europeans | Sweden, UK | 66 | 60,00 | 6,00 | 0,09 | 0,39 |
|  | Central Europeans | Germany, Poland,<br>Slovakia | 90 | 51,00 | 17,00 | 0,11 | 0,42 |
|  | North Balkans | Hungary, Slovenia,<br>Romania | 102 | 46,00 | 19,00 | 0,15 | 0,41 |
|  | Central Balkans | Montenegro,<br>Bosnia and<br>Herzegovina,<br>Kosovo | 160 | 45,00 | 14,00 | 0,11 | 0,45 |
|  | South Balkans | Greece, Macedonia,<br>Bulgaria | 118 | 41,00 | 23,00 | 0,06 | 0,36 |
| West Asia | Iranians | Iran | 92 | 35,00 | 49,00 | 0,08 | 0,47 |
|  | Anatolian and Levant populations | Turkey, Cyprus,<br>Syria, Lebanon | 114 | 35,00 | 35,00 | 0,11 | 0,54 |
|  | Israel populations | Israel | 312 | 32,00 | 35,00 | 0,12 | 0,43 |
|  | Arabs | Saudi Arabia,<br>Yemen, Jordan | 100 | 24,00 | 42,00 | 0,10 | 0,38 |
| Africa | Pygmies (Biaka, Mbuti) | Central African<br>Republic, Congo | 68 | 3,00 | 24,00 | 0,00 | 0,53 |
|  | Ethiopians | Ethiopia | 38 | 9,00 | 39,00 | 0,08 | 0,50 |
|  | West Africans | Nigeria, Senegal | 86 | 10,00 | -4,00 | 0,00 | 0,43 |
|  | South and East Africabs | Botswana, Kenya | 48 | -11,00 | 29,00 | 0,00 | 0,54 |
|  | North Africans | Algeria, Morocco,<br>Egypt | 102 | 32,00 | 9,00 | 0,07 | 0,26 |
| America | Native Americans from South America | Brazil | 58 | -5,00 | -37,00 | 0,00 | 0,00 |
|  | NativeAmericans from Canada and USA | Canada, USA | 50 | 52,00 | -111,00 | 0,02 | 0,36 |
|  | Native Americans from Mexica | Mexico | 98 | 23,00 | -105,00 | 0,04 | 0,05 |
|  | Greenlanders | Denmark | 78 | 64,00 | -43,00 | 0,01 | 0,41 |
| East and<br>South-East<br>Asia | Han | China | 88 | 32,00 | 114,00 | 0,00 | 0,41 |
|  | North China | China | 148 | 43,00 | 109,00 | 0,01 | 0,47 |
|  | South China | China | 116 | 26,00 | 106,00 | 0,00 | 0,41 |
|  | Japanese | Japan | 56 | 38,00 | 138,00 | 0,00 | 0,32 |
|  | South-East Asians | Myanmar,<br>Cambodia | 50 | 16,00 | 101,00 | 0,14 | 0,29 |
|  | Melanesians | Bougainville | 20 | -6,00 | 156,00 | 0,00 | 0,20 |
|  | Papuans | Papua New Guinea | 34 | -6,00 | 144,00 | 0,35 | 0,21 |

### Compiling the dataset on the world populations

In addition to the dataset on the Russian populations, we compiled a dataset on the other regions of the globe. The data came from the 16 papers [14] - [29] which studied indigenous populations by using genome-wide Illumina arrays (Illumina700k, Illumina730k, Illumina660k, Illumina650k, Illumina610k, Illumina550k, and Illumina1M). The resulting merged world dataset included 3336 samples, genotyped by *rs657152* and 66 SNPs in the vicinity of *rs11385942*.

Similar to the Russian dataset we pooled some population samples to achieve larger sample sizes. In most cases we merged samples which came from the same country. Sometimes we have to merge data on the neighbor countries, for example data from Spain and France were merged. Two large countries – China and India – were present by 3 merged populations each. The samples from Russia and neighboring counties present in the world dataset were ignored to avoid double counting the same populations/samples. In total, the world dataset included 32 populations with the average sample size 104 chromosomes. Table 1 presents the list of the analyzed populations (“world dataset” section), their geographic locations are shown on the Figure 1.

### Imputing

One of the two Covid-associated markers, *rs657152*, was directly genotyped in both, Russian and world datasets. The frequencies of this marker in all populations were counted using PLINK software and are present in the Table 1. The second marker, *rs11385942*, is missed in all genome-wide panels available to date. The initial genome-wide association study on COVID-19 [7] genotyped 712,189 SNPs, then imputed 170 million SNPs, and one out of these imputed SNPs, namely *rs11385942*0, demonstrated the highest association with severe COVID-19. Actually there is a haplotype block (approx. 50 kb length [8]) in strong LD, and *rs11385942*0 marks this block nearby as effective as any other SNP in the vicinity. In our study, for the Russian dataset we used a few times more dense panel (4,5 million SNPs) than the panel in [7]. Our panel included 250 SNPs in the vicinity of *rs11385942*0. Imputing was done by the Beagle software [30] by 200 iterations, using 1000 Genomes project dataset as a reference.

As the world dataset was based on the regular (less dense) Illumina panels, including 66 SNPs in the vicinity of *rs11385942*0, we first estimated the imputation quality. For this aim, we compared the genotypes generated for the same samples by using 250 SNPs and 66 SNPs. 1871 out of 1883 genotypes (99,4%) coincided, indicating that regular Illumina panels work well for imputing *rs11385942*0 genotypes. These genotypes were used to count the allele frequencies of *rs11385942*0 in the populations from the world dataset (Table 1).

### Cartographic analysis

The maps of the geographic distribution of the two COVID-associated markers were constructed using the GeneGeo software [31], [32]. The maps of North Eurasia (Russia and neighboring counties) were created with the generalized Shepard’s method, the degree of weight function set to 3 and radius of influence of 1,500 km. The world maps were created by the same method with radius enlarged up to 2,500 km (to fill in gaps between studied populations) and the degree of weight function set to 2 to generate a smoother surface, i.e. trend [33]).

### Statistical analysis

To provide correlation analysis we searched for the number of cases of COVID-19 per 1 M population, recovery rate per 1 M population and mortality caused by this disease per 1 M population, and mortality per finished COVID-19 cases (last updated on September 18, 2020). Because genetic data in each region described the indigenous populations while recovery/mortality rates were available for the total population of a region, we identified 16 regions of Russia and neighboring countries where indigenous populations constitutes the majority (on average, 85% according to the last census) and run correlation analysis on these 16 groups (Table 3). For correlation analysis at the world level we used the same 16 groups from Russian and neighboring countries as well as all groups from the world dataset except data on Native Americans and Greenlanders (Table 4).

To estimate the differences in allele frequencies between groups the Fisher’s exact test was used using the application of GraphPad InStat software package (GraphPad Software, San Diego, CA, USA); the differences were considered statistically significant at p<0.05 with the Bonferroni correction. To determine the correlation between the alleles frequencies and epidemiological parameters per 1 million the Pearson correlation coefficient was calculated using Statsoft Statistica (Dell Statistica, Tulsa, OK, USA); the differences were considered statistically significant at p<0.05 with the Bonferroni correction.

## Results

### Global variation of rs11385942

This single nucleotide insertion was proved to have strongest association with the severe course of SARS-COVID-19 [7]. To unravel its worldwide variation we genotyped 1883 samples from Russian and neighboring countries, and compiled the dataset on 3088 samples from other regions of the globe. The final worldwide dataset contains frequencies of *rs11385942_GA* in 60 populations (Table 1). Figure 2 presents the map of this marker’s variation worldwide. The highest frequency (20-30%) was found in South Asia, followed by West Asia and Europe (5-15%). This marker is rare or not detected in East Asia, North Asia (Siberia), Native Americans, and in Sub-Saharan Africa, while present in North Africa. The data on South-East Asia and Papua are scarce but indicate that this insertion might be present there at elevated frequencies. Generally, the worldwide spread of this marker follows the “West Eurasian” pattern, well-known in human populations genetics for many other genetic markers, including mitochondrial DNA and Y-chromosomal markers [34], [35]. The South Asian populations have much stronger genetic affinities with West Eurasians than with East Eurasians. The specific feature of *rs11385942* distribution pattern is that its frequency is higher in South Asian part rather than in West Asian/European part of the West Eurasian gene pool.

**Figure 2.**
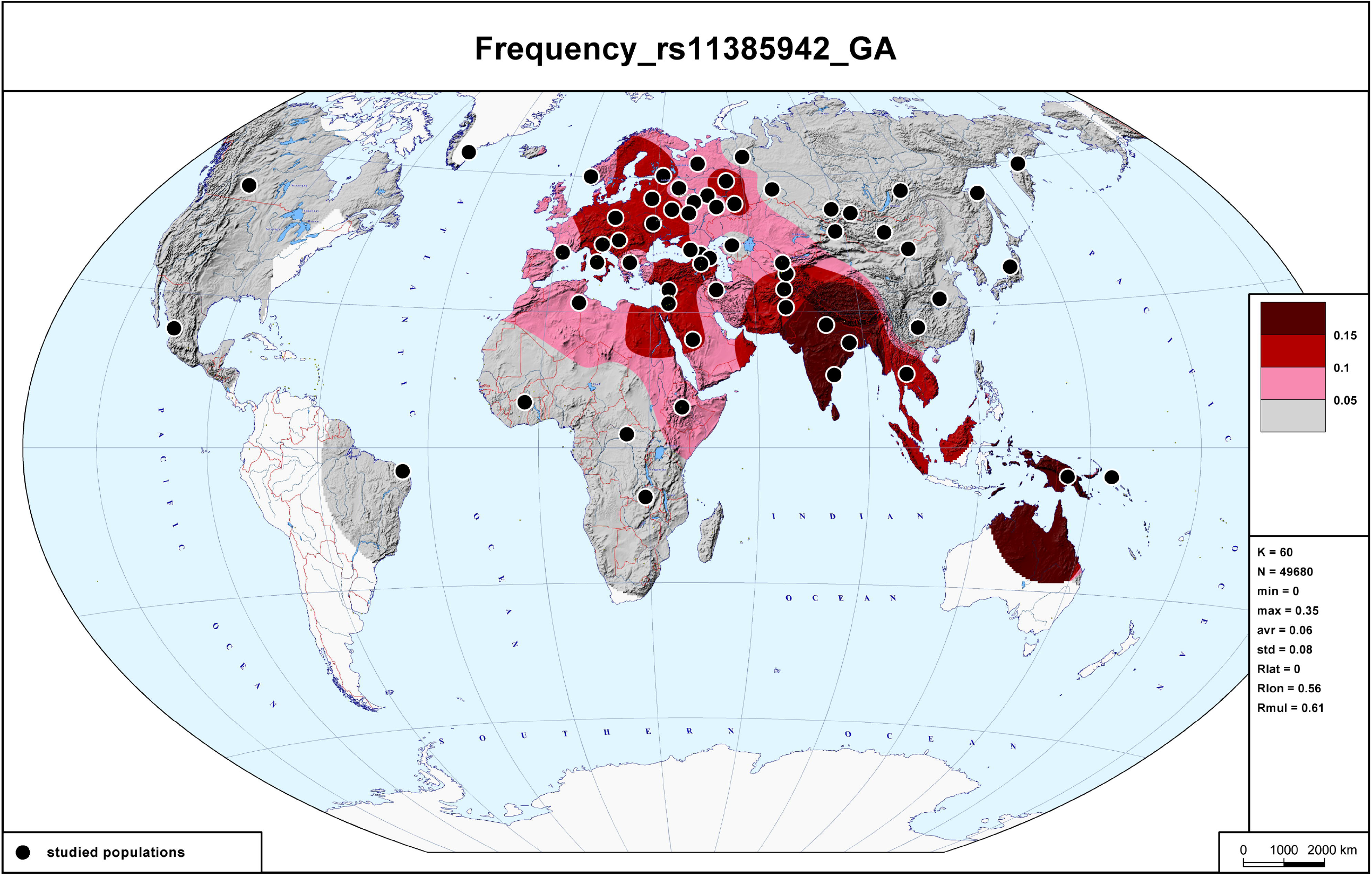
The global variation of *rs11385942_GA* allele. Four colors mark areas with four intervals of frequencies of this risk allele, according to the scale.

This description of a global pattern is inevitable schematic. To compensate this, we coupled the analysis at the worldwide level with the detailed analysis at the national level of the largest country.

### Variation of rs11385942 within Russia

Our extensive Russian dataset allows investigate the variation of *rs11385942* within the area of Russia and neighboring countries in more details. Frequencies are available in the Table 1 and visualized on the map (Figure 3). The map reveals that the difference between higher frequencies in Europe and absence in North Asia (which is obvious on the world map) is not sharp. Instead, there is a pattern of clinal variation, i.e. gradual decrease in frequency throughout 7 thousand kilometers, from the frequencies 13-16% in Ukraine, Belorussia and westernmost Russian populations to the zero frequency in Kamchatka and Chukotka peninsulas at the Pacific coast. Generally, in populations of European Russia the average frequency is 11% while in Siberia it is only 3% (Table 1). In Central Asian countries the frequencies are generally low (1-4%). Tajikistan is the notable exception (frequency is 14%, significant difference from neighbor countries) because Tajikistan population is geographically and genetically close to South Asian populations which exhibit the world maximum of *rs11385942*.

**Figure 3.**
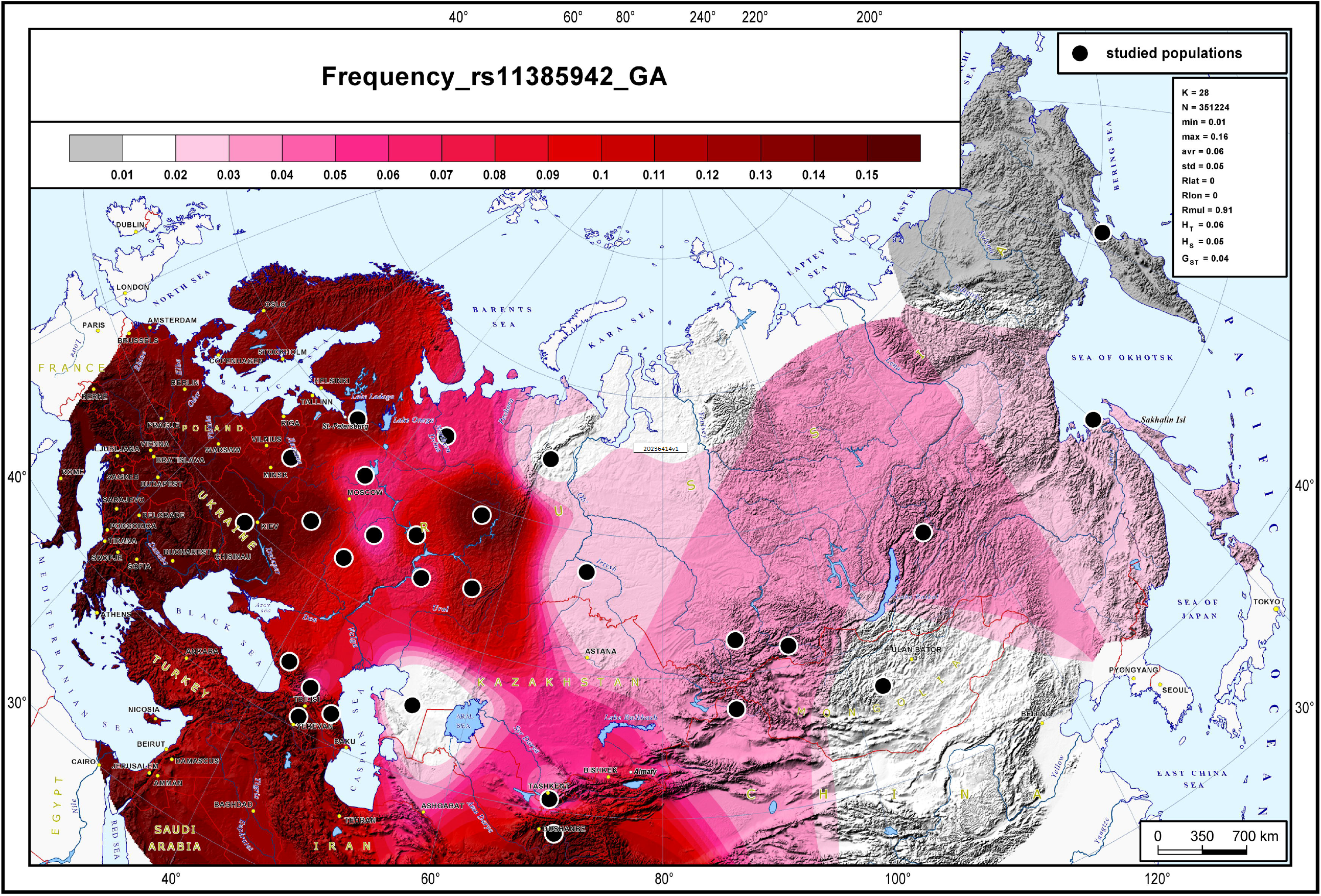
The variation of *rs11385942_GA* within Russia and neighboring countries. The used scale is more detailed that than on the world map (Figure 2).

Focusing on details of the frequency distribution, one may note that *rs11385942* frequency in the southwestern regions of Russia is quite similar in comparison with bordering Belarussia and Ukraine - the differences were insignificant (Table 1). In the Caucasus, the lowest frequency of the risk *GA* allele variant was found in the central regions (Chechnya, Ingushetia, North Ossetia) - 6%, with higher frequency in western and eastern regions (≈ 10%), and particularly in South Caucasus (14%).

### Global variation of rs657152

Figure 4 presents the frequency distribution map of *rs657152*. In comparison with *rs11385942* polymorphic marker, this distribution is more homogeneous. Generally, it is quite frequent among almost all Old World populations. The highest frequencies (above 50%) were observed in Sub-Saharan Africa, while the most Eurasian populations have frequencies between 40 and 50%. At the periphery of Eurasia (Atlantic fridge of Europe, Far East, South-East Asia) the frequency tends to drop below 40% and in Native Americans and Australasians it is nearby absent.

**Figure 4.**
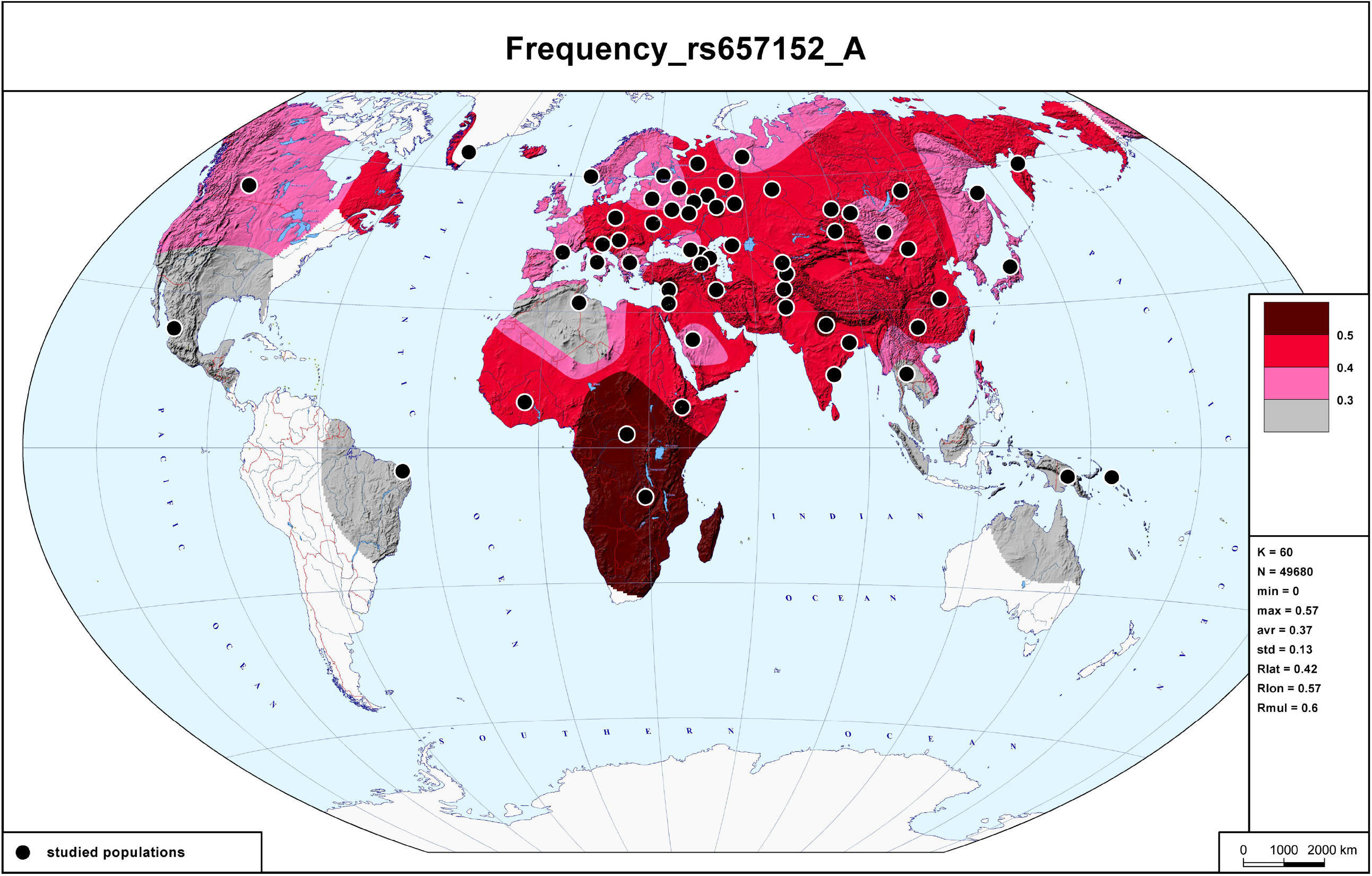
The global variation of *rs657152_A* allele. Four colors mark areas with four intervals of frequencies of this risk allele, according to the scale.

### Variation of rs11385942 within Russia

Among populations of European Russia the *rs657153* frequency varied from 38 to 42%, which was comparable with Belorussia (33%, p>0.05) and lower than in Ukraine (51%, p<0.05) (Tables 1, 2). In Caucasus *rs657152* was most frequently met in eastern regions (Daghestan, 52%), the lowest percentage was found in western regions (27%). Distribution differences in *rs657152* among the population of these regions were most significant in comparison with other included regions (Table 2). In Asia, among Tuvans and Mongols, the frequency of rs657152 was 38-39%, while among the populations of Uzbekistan and Kyrgyzstan the frequency was 51% (with the insignificant differences in comparison with Kazakhstan and Tajikistan, Table 2).

**Table 2.** Pairwise comparison of *rs11385942* polymorphic marker’s *GA* allele variant and *rs657152* polymorphic marker’s *A* allelic variant prevalence in tested populations (Fisher’s exact, p – values with the Bonferroni correction).

| Populations |  |  | (1) | (2) | (3) | (4) | (5) | (6) | (7) | (8) | (9) | (10) | (11) | (12) | (13) | (14) | (15) | (16) |
| --- | --- | --- | --- | --- | --- | --- | --- | --- | --- | --- | --- | --- | --- | --- | --- | --- | --- | --- |
|  |  |  | *rs11385942* |  |  |  |  |  |  |  |  |  |  |  |  |  |  |  |
| Belarus | (1) | *rs657152* |  | 0,4632 | **0,0171** | 0,3622 | 0,7438 | **0,0307** | **0,0132** | 0,5011 | 0,5081 | 0,8775 | 0,4738 | **0,0015** | **0,0001** | **0,0001** | **0,0022** | 0,8765 |
| Ukraine | (2) |  | **0,0008** |  | **0,0001** | **0,0372** | 0,7117 | **0,0002** | **0,0001** | **0,0481** | **0,0412** | 0,5059 | 0,0551 | **0,0001** | **0,0001** | **0,0001** | **0,0001** | 0,5748 |
| Kostroma, Novgorod, Tver, Yaroslavl regions | (3) |  | 0,2290 | **0,0035** |  | 0,1274 | **0,0011** | 0,8676 | 0,8594 | **0,0330** | **0,0275** | **0,0006** | 0,0710 | 0,2395 | **0,0091** | **0,0102** | 0,3816 | **0,0004** |
| Voronezh, Nizhny Novgorod, Ryazan, Tambov regions | (4) |  | 0,1918 | **0,0240** | 0,7849 |  | 0,1313 | 0,1763 | 0,1054 | 0,7702 | 0,7741 | 0,1719 | 0,8720 | **0,0113** | **0,0001** | **0,0002** | **0,0176** | 0,1302 |
| Belgorod, Kaluga, Kursk, Orel, Smolensk regions | (5) |  | 0,3392 | **0,0063** | 0,8540 | 0,6821 |  | **0,0030** | **0,0007** | 0,1712 | 0,1791 | 0,8977 | 0,1823 | **0,0001** | **0,0001** | **0,0001** | **0,0001** | 0,8975 |
| Vologda, Arkhangelsk regions | (6) |  | 0,1181 | **0,0268** | 0,5642 | 0,8512 | 0,5073 |  | 0,7167 | 0,0802 | 0,0663 | **0,0019** | 0,1340 | 0,2231 | **0,0060** | **0,0081** | 0,2798 | **0,0016** |
| Chechnya, Ingushetia, North Ossetia republics | (7) |  | 0,1384 | **0,0336** | 0,6091 | 0,8477 | 0,5593 | 1,0000 |  | **0,0236** | **0,0189** | **0,0007** | 0,0554 | 0,3874 | **0,0335** | **0,0195** | 0,5640 | **0,0006** |
| Dagestan republic | (8) |  | **0,0006** | 0,9366 | **0,0021** | **0,0186** | **0,0047** | **0,0213** | **0,0218** |  | 1,0000 | 0,1859 | 1,0000 | **0,0029** | **0,0001** | **0,0001** | **0,0025** | 0,1771 |
| Karachaevo-Cherkessia and Kabardino-Balkaria republics | (9) |  | 0,1960 | **0,0001** | **0,0004** | **0,0008** | **0,0050** | **0,0001** | **0,0001** | **0,0001** |  | 0,1960 | 0,8887 | **0,0015** | **0,0001** | **0,0001** | **0,0021** | 0,1867 |
| Armenia, Azerbaijan, Georgia | (10) |  | **0,0091** | 0,3363 | 0,0551 | 0,1704 | 0,0651 | 0,2125 | 0,2285 | 0,2621 | **0,0001** |  | 0,2277 | **0,0001** | **0,0001** | **0,0001** | **0,0001** | 1,0000 |
| Chuvashia repu | (11) |  | **0,0275** | 0,2867 | 0,1545 | 0,3205 | 0,1341 | 0,4091 | 0,3965 | 0,2145 | **0,0001** | 0,8577 |  | **0,0074** | **0,0001** | **0,0001** | **0,0082** | 0,1776 |
| Tuva | (12) |  | 0,4102 | **0,0036** | 0,7225 | 0,6174 | 0,9198 | 0,4088 | 0,4500 | **0,0020** | **0,0051** | **0,0402** | 0,0981 |  | 0,3319 | 0,5366 | 0,1504 | **0,0001** |
| Mongolia | (13) |  | 0,2516 | **0,0007** | 0,8853 | 0,6703 | 0,9316 | 0,4480 | 0,4777 | **0,0004** | **0,0002** | **0,0231** | 0,0819 | 0,8685 |  | 0,1958 | 0,8255 | **0,0001** |
| Kazakhstan | (14) |  | **0,0409** | 0,2228 | 0,2206 | 0,4049 | 0,2076 | 0,4984 | 0,4876 | 0,1882 | **0,0001** | 0,7049 | 0,9186 | 0,1518 | 0,1549 |  | 0,0625 | **0,0001** |
| Uzbekistan and Kyrgyzstan | (15) |  | **0,0013** | 0,8743 | **0,0059** | **0,0377** | **0,0108** | **0,0405** | **0,0422** | 0,7517 | **0,0001** | 0,4242 | 0,3311 | **0,0049** | **0,0015** | 0,3019 |  | **0,0001** |
| Tajikistan | (16) |  | **0,0211** | 0,1928 | 0,1428 | 0,3084 | 0,1350 | 0,3984 | 0,3836 | 0,1431 | **0,0001** | 0,7419 | 1,0000 | 0,0863 | 0,0726 | 1,0000 | 0,2556 |  |

### Correlation between frequencies of COVID-associated markers and epidemiological parameters

We compared frequencies of both markers in various populations of Russia with the recovery and mortality rates from COVID-19 in the same populations. The epidemiological numbers (number of COVID-19 cases, recoveries and death) and population sizes are present in the Table 3. We have observed (Table 5) a negative insignificant correlations between the frequency of both risk polymorphic markers and the parameters linked with the number of cases of COVID-19 (either absolute numbers or numbers normalized by the population size). Instead, we found positive correlations between both risk alleles and the mortality rate, measured as number of deaths per number of Covid-19 cases, and negative correlation between both risk alleles and the recovery rate (number of recoveries normalized by number of cases). The correlation of mortality with the frequency of *rs657152* (r=0,63) was significant with p-value 0,01 (Table 5), while correlation with the frequency of *rs11385942* (r=0,24) was insignificant (p=0.38). Correlation between *rs657152* and mortality rate remained significant after Bonferroni correction. When analyzed the combined dataset (Russian and world populations), all correlations became insignificant (Table 5).

**Table 3.**
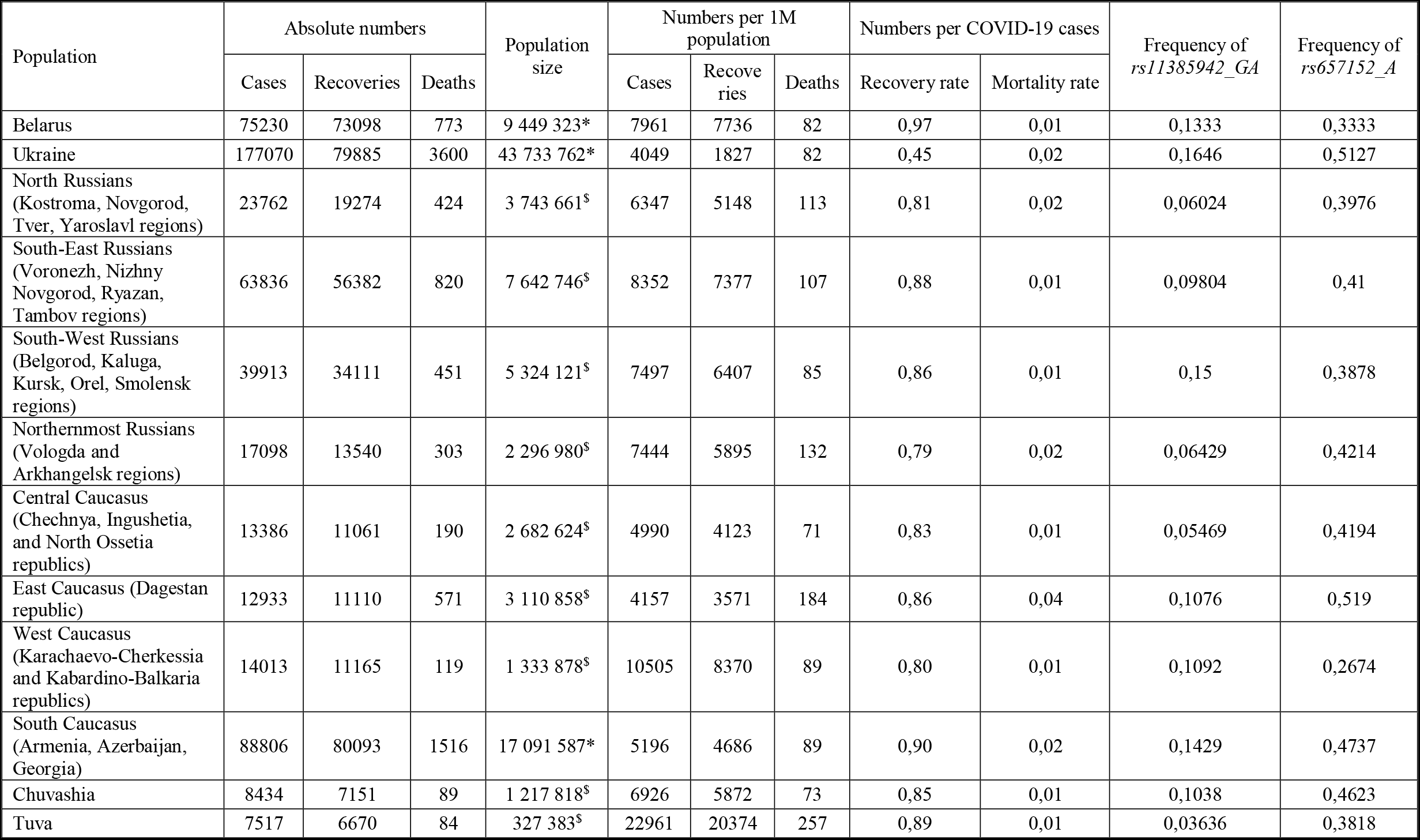
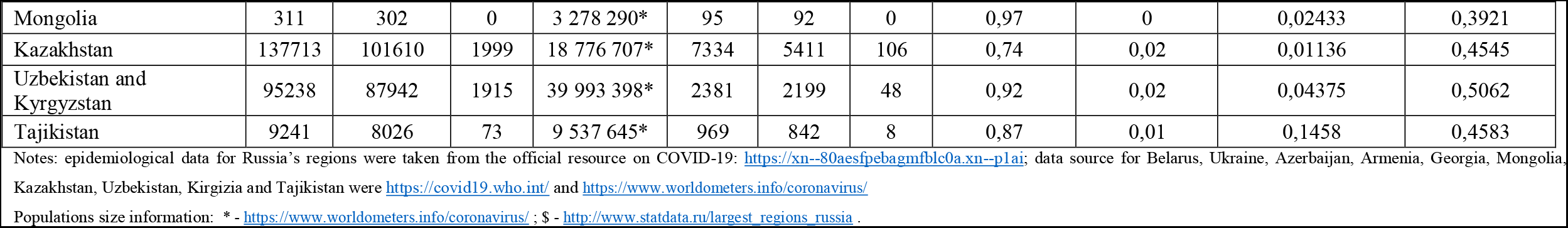
Epidemiological parameters (relevant for September 18, 2020) and frequencies of the two genetic markers associated with the severe COVID-19 (dataset on Russia and neighboring countries).

| Population | Absolute numbers |  |  | Population size | Numbers per 1M population |  |  | Numbers per COVID-19 cases |  | Frequency of *rs11385942_GA* | Frequency of *rs657152_A* |
| --- | --- | --- | --- | --- | --- | --- | --- | --- | --- | --- | --- |
|  | Cases | Recoveries | Deaths |  | Cases | Recoveries | Deaths | Recovery rate | Mortality rate |  |  |
| Belarus | 75230 | 73098 | 773 | 9 449 323* | 7961 | 7736 | 82 | 0,97 | 0,01 | 0,1333 | 0,3333 |
| Ukraine | 177070 | 79885 | 3600 | 43 733 762* | 4049 | 1827 | 82 | 0,45 | 0,02 | 0,1646 | 0,5127 |
| North Russians<br>(Kostroma, Novgorod, Tver, Yaroslavl regions) | 23762 | 19274 | 424 | 3 743 661<sup>\$</sup> | 6347 | 5148 | 113 | 0,81 | 0,02 | 0,06024 | 0,3976 |
| South-East Russians<br>(Voronezh, Nizhny Novgorod, Ryazan, Tambov regions) | 63836 | 56382 | 820 | 7 642 746<sup>\$</sup> | 8352 | 7377 | 107 | 0,88 | 0,01 | 0,09804 | 0,41 |
| South-West Russians<br>(Belgorod, Kaluga, Kursk, Orel, Smolensk regions) | 39913 | 34111 | 451 | 5 324 121<sup>\$</sup> | 7497 | 6407 | 85 | 0,86 | 0,01 | 0,15 | 0,3878 |
| Northernmost Russians<br>(Vologda and Arkhangelsk regions) | 17098 | 13540 | 303 | 2 296 980<sup>\$</sup> | 7444 | 5895 | 132 | 0,79 | 0,02 | 0,06429 | 0,4214 |
| Central Caucasus<br>(Chechnya, Ingushetia, and North Ossetia republics) | 13386 | 11061 | 190 | 2 682 624<sup>\$</sup> | 4990 | 4123 | 71 | 0,83 | 0,01 | 0,05469 | 0,4194 |
| East Caucasus (Dagestan republic) | 12933 | 11110 | 571 | 3 110 858<sup>\$</sup> | 4157 | 3571 | 184 | 0,86 | 0,04 | 0,1076 | 0,519 |
| West Caucasus<br>(Karachaevo-Cherkessia and Kabardino-Balkaria republics) | 14013 | 11165 | 119 | 1 333 878<sup>\$</sup> | 10505 | 8370 | 89 | 0,80 | 0,01 | 0,1092 | 0,2674 |
| South Caucasus<br>(Armenia, Azerbaijan, Georgia) | 88806 | 80093 | 1516 | 17 091 587* | 5196 | 4686 | 89 | 0,90 | 0,02 | 0,1429 | 0,4737 |
| Chuvashia | 8434 | 7151 | 89 | 1 217 818<sup>\$</sup> | 6926 | 5872 | 73 | 0,85 | 0,01 | 0,1038 | 0,4623 |
| Tuva | 7517 | 6670 | 84 | 327 383<sup>\$</sup> | 22961 | 20374 | 257 | 0,89 | 0,01 | 0,03636 | 0,3818 |
| Mongolia | 311 | 302 | 0 | 3 278 290* | 95 | 92 | 0 | 0,97 | 0 | 0,02433 | 0,3921 |
| Kazakhstan | 137713 | 101610 | 1999 | 18 776 707* | 7334 | 5411 | 106 | 0,74 | 0,02 | 0,01136 | 0,4545 |
| Uzbekistan and<br>Kyrgyzstan | 95238 | 87942 | 1915 | 39 993 398* | 2381 | 2199 | 48 | 0,92 | 0,02 | 0,04375 | 0,5062 |
| Tajikistan | 9241 | 8026 | 73 | 9 537 645* | 969 | 842 | 8 | 0,87 | 0,01 | 0,1458 | 0,4583 |

Notes: epidemiological data for Russia's regions were taken from the official resource on COVID-19: https://xn--80aesfpebagmblc0a.xn--p1ai; data source for Belarus, Ukraine, Azerbaijan, Armenia, Georgia, Mongolia,
Kazakhstan, Uzbekistan, Kirgizia and Tajikistan were https://covid19.who.int/ and https://www.worldometers.info/coronavirus/
Populations size information: \* - https://www.worldometers.info/coronavirus/ ; \$ - http://www.statdata.ru/largest_regions_russia .

**Table 4.**
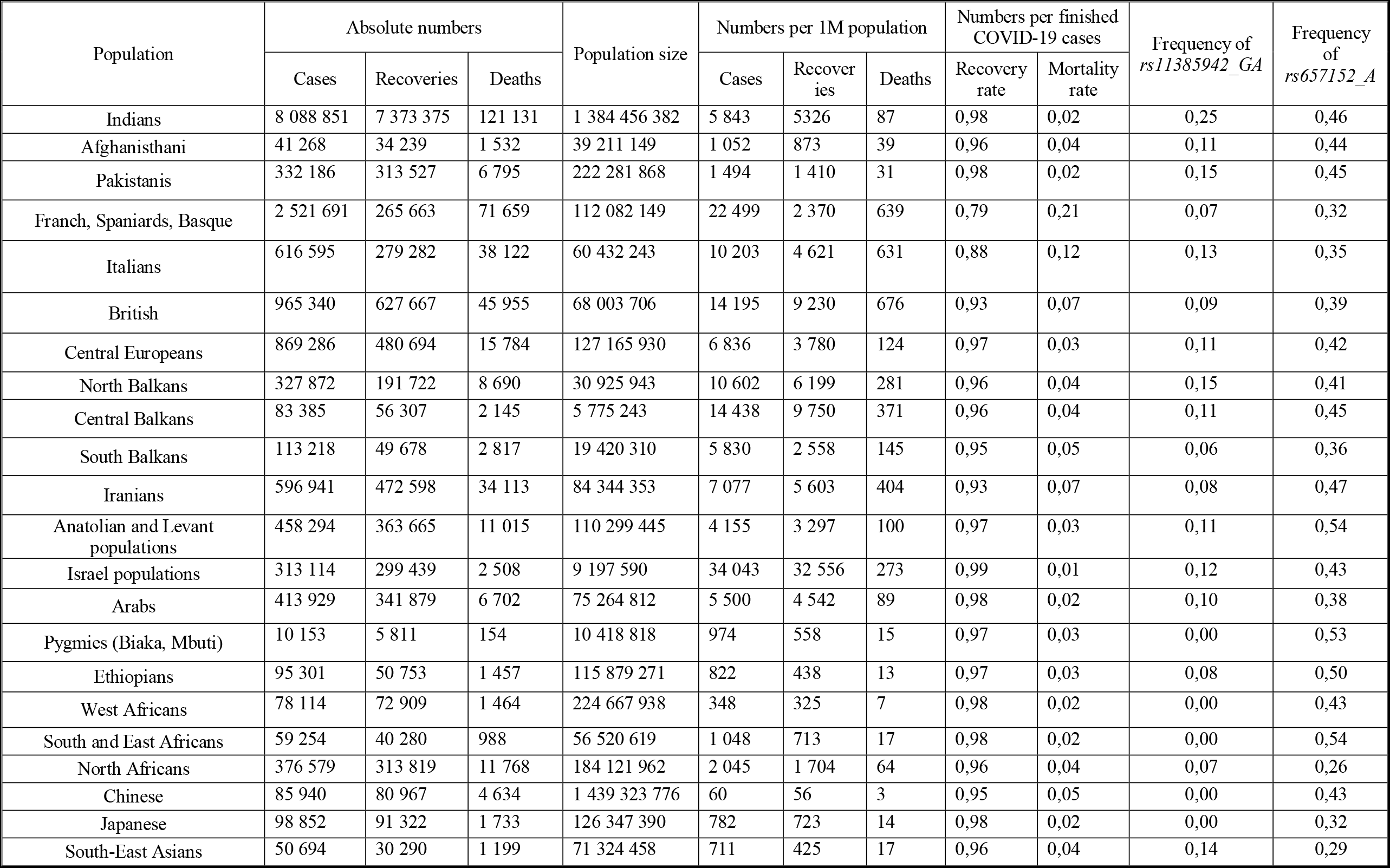
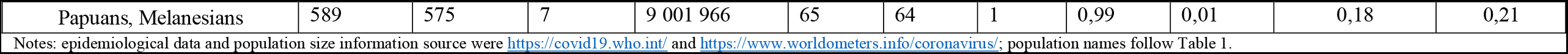
COVID-19 epidemiological parameters and allele frequencies (dataset on other countries).

| Population | Absolute numbers |  |  | Population size | Numbers per 1M population |  |  | Numbers per finished COVID-19 cases |  | Frequency of *rs11385942_GA* | Frequency of *rs657152_A* |
| --- | --- | --- | --- | --- | --- | --- | --- | --- | --- | --- | --- |
|  | Cases | Recoveries | Deaths |  | Cases | Recoveries | Deaths | Recovery rate | Mortality rate |  |  |
| Indians | 8 088 851 | 7 373 375 | 121 131 | 1 384 456 382 | 5 843 | 5326 | 87 | 0,98 | 0,02 | 0,25 | 0,46 |
| Afghanisthani | 41 268 | 34 239 | 1 532 | 39 211 149 | 1 052 | 873 | 39 | 0,96 | 0,04 | 0,11 | 0,44 |
| Pakistanis | 332 186 | 313 527 | 6 795 | 222 281 868 | 1 494 | 1 410 | 31 | 0,98 | 0,02 | 0,15 | 0,45 |
| Franch, Spaniards, Basque | 2 521 691 | 265 663 | 71 659 | 112 082 149 | 22 499 | 2 370 | 639 | 0,79 | 0,21 | 0,07 | 0,32 |
| Italians | 616 595 | 279 282 | 38 122 | 60 432 243 | 10 203 | 4 621 | 631 | 0,88 | 0,12 | 0,13 | 0,35 |
| British | 965 340 | 627 667 | 45 955 | 68 003 706 | 14 195 | 9 230 | 676 | 0,93 | 0,07 | 0,09 | 0,39 |
| Central Europeans | 869 286 | 480 694 | 15 784 | 127 165 930 | 6 836 | 3 780 | 124 | 0,97 | 0,03 | 0,11 | 0,42 |
| North Balkans | 327 872 | 191 722 | 8 690 | 30 925 943 | 10 602 | 6 199 | 281 | 0,96 | 0,04 | 0,15 | 0,41 |
| Central Balkans | 83 385 | 56 307 | 2 145 | 5 775 243 | 14 438 | 9 750 | 371 | 0,96 | 0,04 | 0,11 | 0,45 |
| South Balkans | 113 218 | 49 678 | 2 817 | 19 420 310 | 5 830 | 2 558 | 145 | 0,95 | 0,05 | 0,06 | 0,36 |
| Iranians | 596 941 | 472 598 | 34 113 | 84 344 353 | 7 077 | 5 603 | 404 | 0,93 | 0,07 | 0,08 | 0,47 |
| Anatolian and Levant populations | 458 294 | 363 665 | 11 015 | 110 299 445 | 4 155 | 3 297 | 100 | 0,97 | 0,03 | 0,11 | 0,54 |
| Israel populations | 313 114 | 299 439 | 2 508 | 9 197 590 | 34 043 | 32 556 | 273 | 0,99 | 0,01 | 0,12 | 0,43 |
| Arabs | 413 929 | 341 879 | 6 702 | 75 264 812 | 5 500 | 4 542 | 89 | 0,98 | 0,02 | 0,10 | 0,38 |
| Pygmies (Biaka, Mbuti) | 10 153 | 5 811 | 154 | 10 418 818 | 974 | 558 | 15 | 0,97 | 0,03 | 0,00 | 0,53 |
| Ethiopians | 95 301 | 50 753 | 1 457 | 115 879 271 | 822 | 438 | 13 | 0,97 | 0,03 | 0,08 | 0,50 |
| West Africans | 78 114 | 72 909 | 1 464 | 224 667 938 | 348 | 325 | 7 | 0,98 | 0,02 | 0,00 | 0,43 |
| South and East Africans | 59 254 | 40 280 | 988 | 56 520 619 | 1 048 | 713 | 17 | 0,98 | 0,02 | 0,00 | 0,54 |
| North Africans | 376 579 | 313 819 | 11 768 | 184 121 962 | 2 045 | 1 704 | 64 | 0,96 | 0,04 | 0,07 | 0,26 |
| Chinese | 85 940 | 80 967 | 4 634 | 1 439 323 776 | 60 | 56 | 3 | 0,95 | 0,05 | 0,00 | 0,43 |
| Japanese | 98 852 | 91 322 | 1 733 | 126 347 390 | 782 | 723 | 14 | 0,98 | 0,02 | 0,00 | 0,32 |
| South-East Asians | 50 694 | 30 290 | 1 199 | 71 324 458 | 711 | 425 | 17 | 0,96 | 0,04 | 0,14 | 0,29 |
| Papuans, Melanesians | 589 | 575 | 7 | 9 001 966 | 65 | 64 | 1 | 0,99 | 0,01 | 0,18 | 0,21 |

Notes: epidemiological data and population size information source were https://covid19.who.int/ and https://www.worldometers.info/coronavirus/; population names follow Table 1.

**Table 5.** Correlations between recoveries, mortalities and frequencies of genetic markers

| Epidemiological parameter | “Russian dataset” |  | “World dataset” |  |
| --- | --- | --- | --- | --- |
|  | rs11385942_GA | rs657152_A | rs11385942_GA | rs657152_A |
| Number of COVID-19 cases per 1 million persons | -0,18 (p=0,50) | -0,44 (p=0,09) | 0,11 (p=0,49) | -0,13 (p=0,44) |
| Number of recoveries per 1 million persons | -0,18 (p=0,50) | -0,46 (p=0,08) | 0,13 (p=0,43) | -0,03 (p=0,84) |
| Number of deaths per 1 million persons | -0,17 (p=0,52) | -0,04 (p=0,89) | 0,10 (p=0,54) | -0,12 (p=0,45) |
| Mortality rate (number of deaths per finished COVID-19 cases) | 0,24 (p=0,38) | **0,63 (p=0,01)** | -0,03 (p=0,88) | -0,14 (p=0,38) |

Notes: p-values are indicated in the parenthesis

## Discussion

The genome-wide association study by Ellinghaus D. et al. revealed the strongest association signal at locus 3p21.31 comprised six genes (including SLC6A20, LZTFL1, CCR9, FYCO1, CXCR6, XCR1): *rs11385942* risk allele *GA* is associated with a genetic predisposition to COVID-19 mediated acute respiratory failure. The frequency of risk allele was higher among patients who received mechanical ventilation compared to those who received only oxygen supplementation. The most relevant genes to COVID-19 were SLC6A20, LZTFL1 and CXCR6. The gene SLC6A20 encodes sodium-iminoacid transporter 1, which closely interacts with ACE2 – the “entrance gate” of SARS-CoV-2 [36], [37]. LZTFL1 regulates the functions of cilium and intra-flagellar transport in cells [38]. The CXCR6 gene regulates the specific location of resident memory T cells in different parts of the lung, providing a stable immune response to pathogens in respiratory tract [39]. The carriage of *GA* risk allele of determined the decreased expression of CXCR6 and the increased expression of SLC6A20 and LZTFL1 genes in human lung cells, which determines the severe COVID-19 [7].

Ellinghaus D. et al noted that the frequency of risk alleles *rs11385942* varies among populations worldwide, but did not analyze the pattern of this variation. We generated the dataset on the frequencies of this marker in various world populations and identified that its global variation follows the West Eurasian pattern. Like many other polymorphisms, *rs11385942* is frequent in West Eurasians (Europeans, West Asians, and South Asians) but rare or absent in all other parts of the globe, including East Asians, North Asians, Native Americans and Sub-Saharan Africans (North African populations are related to West Eurasians and thus carry this markers at moderate frequencies). This marker (along with the linked SNPs in the 50kb length haplotype block) came from admixture with Neanderthals [8]. This explains well its absence in Sub-Saharan Africa (where no Neanderthals are known), while absence in East Eurasia can be attributed to genetic drift after separating West and East Eurasians. Our detailed analysis of populations from Russia demonstrated the absence of abrupt differences between Eurasian subcontinents: for example, elevated frequency of *rs11385942* in Europe and absence in the Pacific coast are linked by the chain of populations with intermediate frequencies, gradually decreasing eastward.

The genome site exhibiting the second high association with severe COVID-19, *rs657152* is located within the AB0 blood group locus. The importance of AB0 blood groups system for COVID-19 was also reported in other studies, namely the risk of respiratory failure in COVID-19 was highest in patients with blood group A compared to other groups while the lowest risk was in patients with blood group O, [40], [41], though these studies are primarily concerned with the risk of infection and not with disease severity [42]. The protective effect of blood group 0, in contrast to other groups, was explained by the presence of neutralizing antibodies against protein-linked N-glycans [7], [43]. It is also known that there is a linkage between ABO blood group locus and the expression of the von Willebrand factor’s gene (VWF) (12p13.31 locus), which in connection with VIII factor promotes the clots formation on the surface of damaged vessels. Pulmonary endothelial cells in non-O groups are associated with higher levels of VWF compared to O group [44], [45], which may explain the role of AB0 blood group locus in COVID-19 patients.

However, the *rs657152* does not directly encode blood group A or any other blood group. Instead, it can be used to distinguish rare variants [46]. The fact that GWAS identified the strongest association with this SNP which does not code the blood group while other studies identified association with the classical blood groups indicates the complexity underlying these associations which might be resolved by future studies involving the sequencing of the entire AB0 locus. Meanwhile, we can consider the frequency distribution of AB0 group A (Figure 5). The most impressive feature of this map is the large amount of data it is based on: 2757 populations, which is almost thirty times more than the dataset for the other SNPs analyzed in this study. This was because the map (Figure 5) is based on the frequencies of blood groups accumulated in 20^th^ century due to works of many scholars. This case exemplifies the value of old-fashion datasets, because using modern datasets (like 1000 Genomes Project or the dataset analyzed in our study) one could obtain a rough, approximate distribution map, while using the data from books published in the second half of 20^th^ century provided the very detailed and reliable picture (Figure 5). This picture demonstrated that highest frequencies (30% and higher) were observed in West Europe, slightly lower in the Volga-Ural region in East Europe, and generally low frequencies (10-20%) in North Asia (Siberia), East Asia, and Sub-Saharan Africa. As for South Asian populations, most of them carry AB0_A at moderate frequencies (15-20%) while neighboring West Central Asians exhibit almost as high frequencies, as West Europeans do. Summarizing, we observed the higher frequency of the first risk allele *rs11385942* in South Asia, somewhat lower in Europe and West Asia, and low in other regions of the globe. For the second risk allele *rs657152* we observed the highest frequency in Africa and moderate in Eurasia. Finally, the AB0 group A is most frequent in Europe and moderately frequent across Eurasia. So that, Europeans and South Asians carry every risk allele at highest or high frequencies, as compared with populations from other regions.

**Figure 5.**
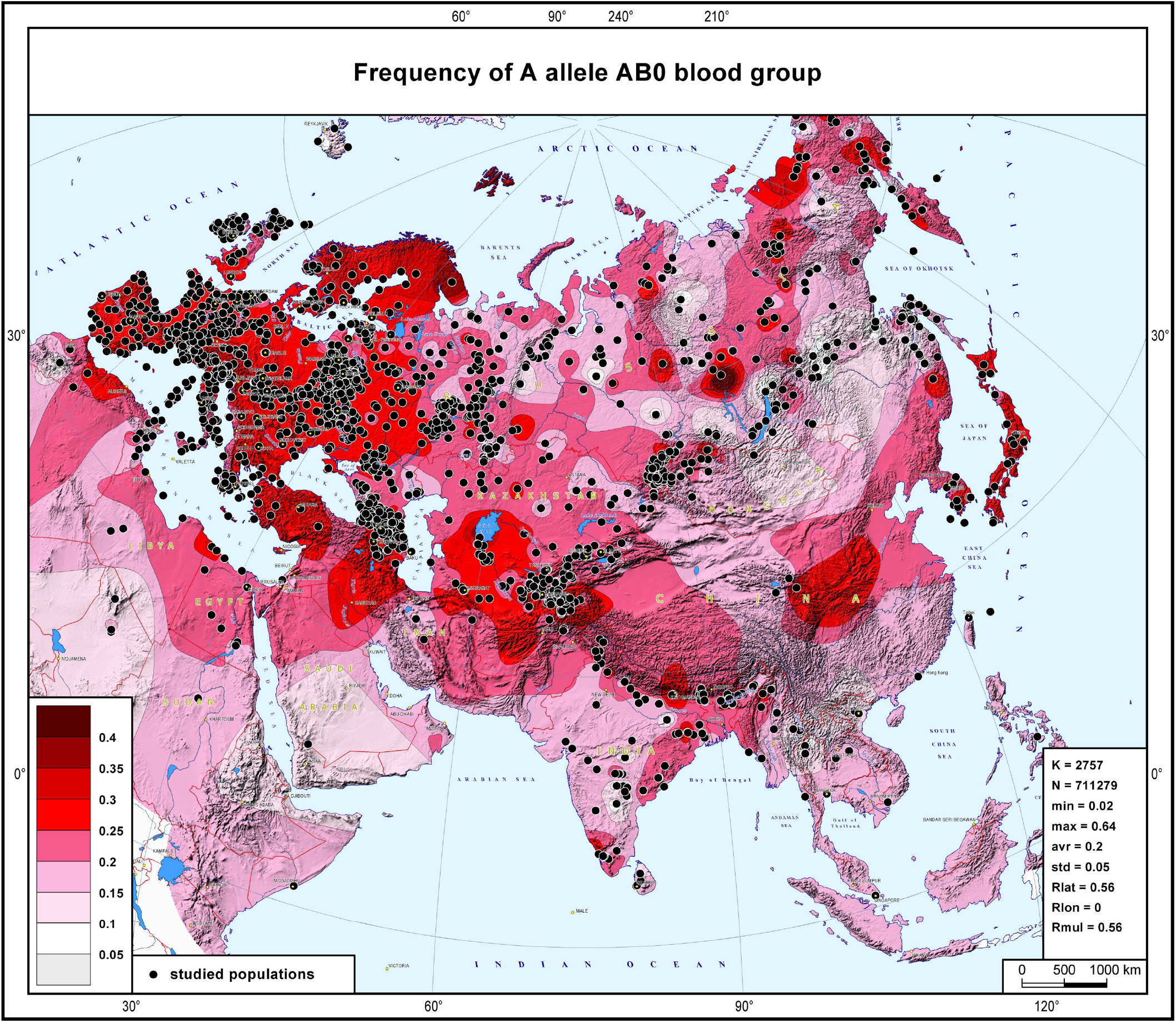
The global variation of blood group A (AB0 system). The map was modified from [47]

Therefore, we investigated whether the elevated frequency of risk alleles in a population indeed results in worse epidemiological situation in this country or region. Though we cannot claim the causative effect, we found the reasonable pattern of correlations. First, we found no correlation between frequencies of risk alleles and numbers of COVID-19 cases: this was expected because these alleles are associated with severe course of disease rather than with the susceptibility. Second, the correlations with the outcomes of the disease were as they should be: for both markers, higher population frequency of risk alleles correlated positively with the deaths rate. For the *rs11385942* we can state the tendency only, while for *rs657152* the correlation was high (r=0,6) and significant (p-value 0,01, Table 5). Notably, we found these reasonable correlations on the “Russian” dataset only, while correlations on the world dataset were zero. This difference could be attributed to the pronounced difference between countries how the not-severe cases are counted: the number of underestimated COVID-19 cases affects the mortality rate dramatically, making it impossible to reveal the correlation with genetic factors. On the contrary, when using epidemiological statistics within one country (in our case, Russia) the ways to count COVID-19 cases become more uniform, and the correlation with variation of the risk alleles frequency becomes evident. The genetic data (allele frequencies) used in our study are also affected by relatively low samples sizes, thus statistical noise is expected to decrease the correlation values. To this end we believe that future studies will identify even stronger correlation between recovery/mortality rates and population’s gene pool, and that genetic variation between populations makes small but real contribution into the heterogeneity of the pandemic worldwide.

## Data Availability

Authors declare that all data provided in this manuscript is available and could be submitted on request.

